# Acute exercise induces natural killer cell mobilization but not infiltration into prostate cancer tissue in humans

**DOI:** 10.1101/2022.04.28.22274307

**Authors:** A Schenk, T Esser, S Belen, N Gunasekara, N Joisten, M Winker, L Weike, W Bloch, A Heidenreich, J Herden, H Löser, S Oganesian, S Theurich, C Watzl, P Zimmer

## Abstract

The mobilization and activation of natural killer (NK) cells have been proposed as key mechanisms promoting anti oncogenic effects of physical exercise. Although mouse models have proven that physical exercise recruits NK cells to tumor tissue and inhibits tumor growth, this preclinical finding has not been transferred to the clinical setting yet. In this first-in-human study, we found that physical exercise mobilizes and redistributes NK cells, especially those with a cytotoxic phenotype, in line with preclinical models. However, physical exercise did not increase NK cell tumor infiltrates. Future studies should carefully distinguish between acute and chronic exercise modalities and should be encouraged to investigate more immune responsive tumor entities.

## Manuscript

Physical exercise supports amelioration of side effects arising from cancer treatment and improves patients’ physical function and quality of life (Schmitz et al., 2019). Epidemiological studies further suggest that regular physical activity reduces cancer risk and mortality (Patel et al., 2019). Preclinical studies support these findings and indicate that physical exercise reduces tumor growth in various types of cancer (Eschke et al., 2019). In this context, exercise-induced mobilization and activation of Natural Killer (NK) cells belong to the most highlighted potential underlying mechanisms (Holmen Olofsson et al., 2022). In their spectacular work, Pedersen and colleagues have shown that physical exercise mobilizes NK cells and subsequently increase NK cell infiltrates in B16 tumors in mice, thereby inhibiting disease progression and severity (Pedersen et al., 2016). A recent meta-analysis indicates an anticarcinogenic effect of exercise-conditioned human serum on tumor cell growth in vitro (Orange, Jordan, & Saxton, 2020). However, the impact of exercise on human tumor tissue has not been investigated yet.

Here, we investigated in a first-in-human trial, whether an acute bout of aerobic exercise, which has previously shown to be the most potent stimulus for immune cell and specifically NK cell mobilization (Schlagheck et al., 2020), alters the composition of tumor immune cell infiltrates in patients with prostate cancer undergoing radical prostatectomy. For this purpose, 22 patients were recruited three weeks prior to surgery. Study participants underwent an initial screening, including a cardiorespiratory exercise test (CPET, see methods) and were subsequently randomized into a passive control or an exercise group. The exercise group performed a 30-minute aerobic exercise session at an intensity of 75% peak oxygen consumption the evening (7:00 p.m.) prior to surgery (next morning, 8:00-10:00 a.m.). Blood samples (serum and PBMCs) were collected before (t1) and after (t2) the exercise intervention, as well as immediately before surgery (t3). Dissected tumor tissue was collected for analysis of NK cell infiltrates at t3. The study design and sample characteristics are shown in figure 1 (A, B).

**Fig 1:**
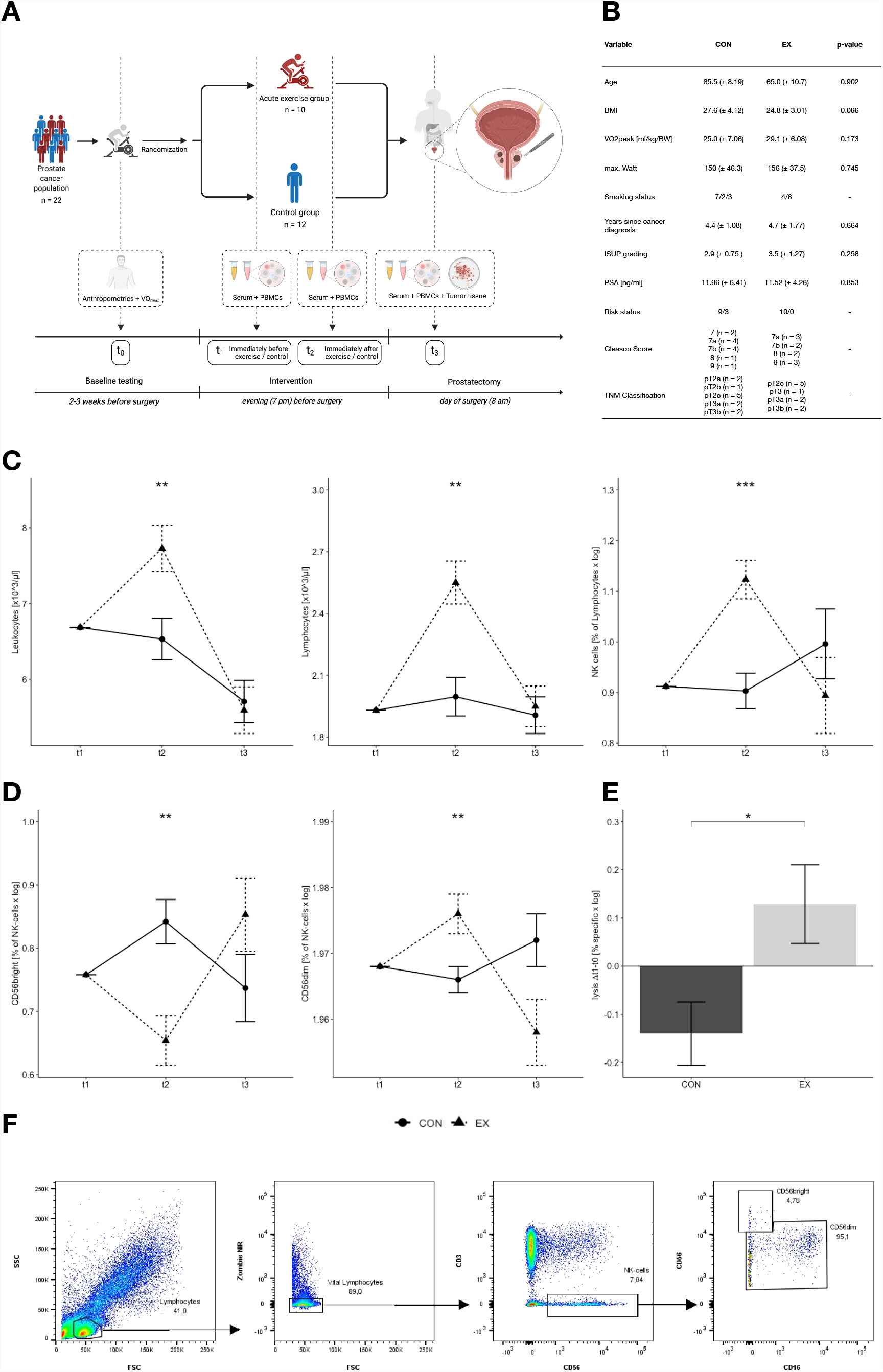
Increased mobilization and characteristics of leukocytes, lymphocytes and NK cells after acute exercise in prostate cancer patients. A) Experimental design. B) Participant characteristics. Smoking status reported as smoker/not-smoker/former smoker. Years since cancer diagnosis refers to the year where the study was conducted (2022). Risk status is reported as high-risk/intermediate-risk. Data is presented as means ± SD. Groups are compared using unpaired t-tests. C) Exercise-induced immune cell changes (CON n = 12, EX n = 10, baseline adjusted repeated measures (t1-t3) ANCOVA). D) Exercise-induced phenotype-specific NK cell changes (CON n = 12, EX n = 10, baseline adjusted repeated measures (t1-t3) ANCOVA). E) NK-cell cytotoxicity displayed as percentage of killed K562 tumor cells in an effector to target ratio of 1:1. Changes in lysis activity were compared using one-way ANCOVA adjusted for changes in CD56^*dim*^/CD56^*bright*^ ratio. (CON n = 12, EX n = 10). F) NK-cell gating. Lymphocytes were gated by size and granularity via forward (FSC) and sideward scatter (SSC). Thereafter, dead lymphocytes were excluded via Zombie NIR staining. NK-cells were gated from the vital lymphocytes as CD3 CD56. Subsequently, NK cells were divided into CD56^*bright*^ and CD56^*dim*^ NK cells according to Montaldo et al. (2013) as CD56^*bright*^ being CD56^*bright*^ CD16 and CD56^*dim*^ being CD56^*dim*^ CD16 /. Data are presented as means ± SEM. Statistical significances are displayed for interactions in ANCOVA and t-test as *p < 0.05, **p < 0.01, ***p < 0.001.

First, we investigated whether acute exercise induces a mobilization of immune cells. In accordance with published data, aerobic exercise increased total numbers of both, circulating leukocytes and lymphocytes, while both return to pre exercise levels before surgery (figure 1C). We next focused on the NK cell compartment, showing a distinct and phenotype-specific mobilization. In line with the literature, the exercise session primarily mobilized NK cells of a cytotoxic phenotype (CD56^dim^), showing the typically pattern of increased proportions immediately after exercise, followed by a rapid decrease which still can be observed the next morning (t3). Cytokine producing CD56^bright^ NK-cells revealed an opposite and less pronounced reaction (figure 1D). Since acute exercise has been described to potentially also improve NK cell function (Rumpf et al., 2021), we conducted flow cytometry-based cytotoxicity assays (see methods). In accordance with a recent meta-analysis (Rumpf et al., 2021) and in contrast to the mouse study of Pedersen et al. (Pedersen et al., 2016), exercise intermediately significantly increases the cytolytic activity of NK cells, even when adjusting for changes in the CD56^dim^/CD56^bright^ ratio (figure 1D).

Pedersen and colleagues reported that especially Interleukin-6 (IL6) sensitive NK cells were redistributed to the tumor tissue (Pedersen et al., 2016). Blocking IL6 led to reduced NK cell infiltrates and enhanced tumor growth in mice (Pedersen et al., 2016). We therefore assessed serum levels of IL6 and other cytokines (Chow et al., 2022). Serum levels of IL6 significantly increase with exercise, although less pronounced compared to the animal model (IL6 increase in mice: 7 fold, IL6 increase in our study: 2 fold) (figure 2A). Of note, internal load of endurance exercise intensity and duration is hardly comparable between caged rodents and humans for several reasons, including different natural physical activity patterns and muscle fibre composition (Joisten, Schenk, & Zimmer, 2020). TNF-α, IL10, and MMP-2 serum levels did not change over time.

**Fig 2:**
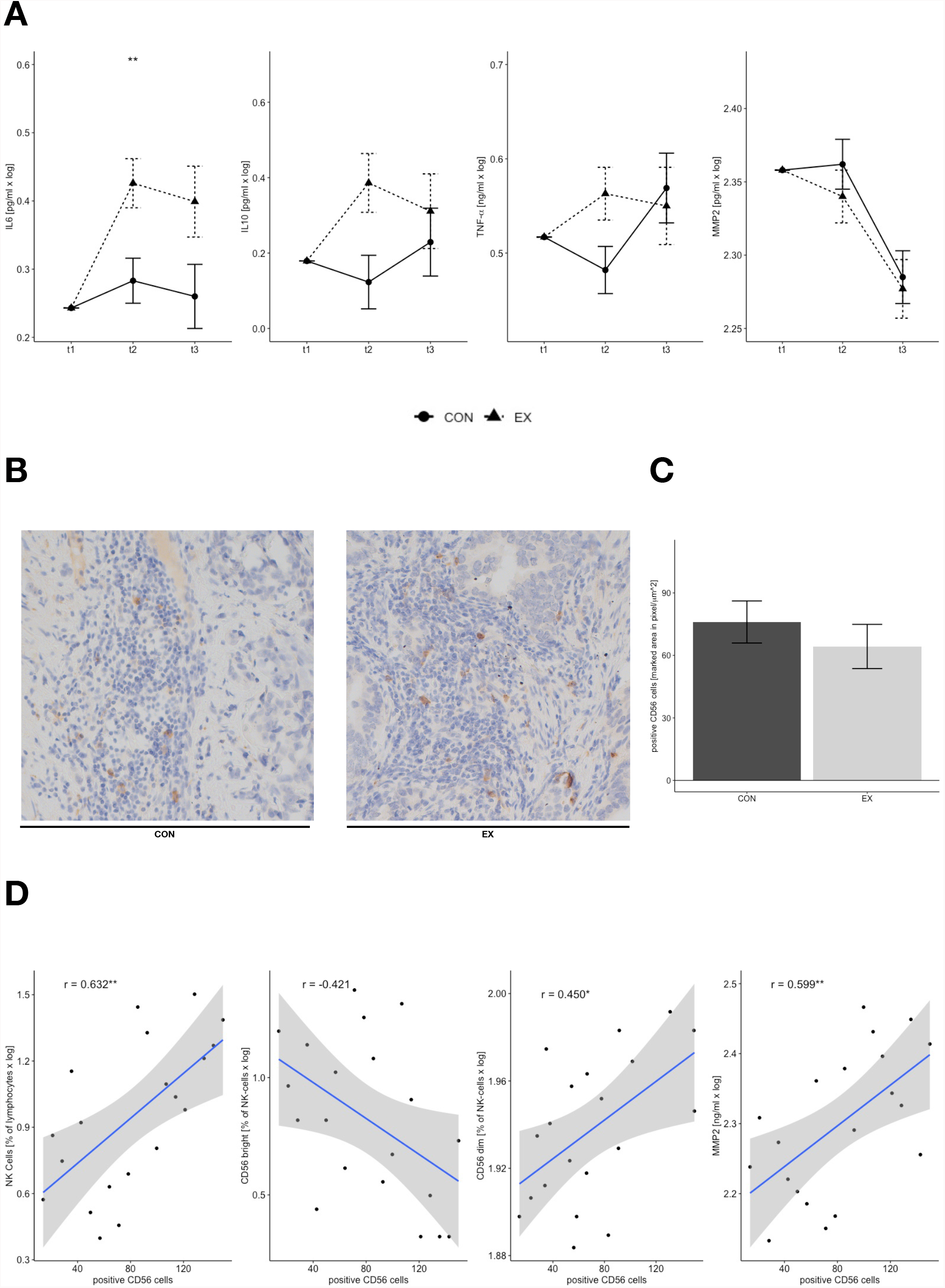
Cytokine and NK cell tumor infiltrates in prostate cancer patients after acute exercise vs. control. A) Exercise-induced cytokine changes in blood serum (CON n = 12, EX n = 10, baseline adjusted repeated measures (t1-t3) ANCOVA). B) Histological analysis of tumor tissue stained for CD56 cells, 20x magnification. C) Quantification of B (CON n = 12, EX n = 10, one-way ANCOVA adjusted for circulation NK cells at t3) D) Spearman rank-correlations between tumor infiltrates and NK cells, NK phenotype and Matrix-metalloproteinase-2 measured in peripheral blood. (n = 22). Data are presented as means ± SEM. Statistical significances are displayed for interactions in ANCOVAs and correlations as *p < 0.05, **p < 0.01, ***p < 0.001.

In contrast to the animal study, we did not observe any differences of NK cell infiltrates in the tumor tissue between the intervention- and the control group (figure 2B and C). However, circulating NK cell numbers which have been assessed immediately prior to surgery were associated with NK cell infiltrates in a phenotype-specific manner (figure 2D). Greater levels of the CD56^dim^ NK cells subset correlated with NK cell tumor infiltrates, whereas the CD56^bright^ NK cell subset was inversely related (figure 2D). In addition, greater MMP-2 levels, a protease which is crucial for NK cell transmigration and which has previously been shown to be influenced through regular exercise, were associated with both, circulating NK cells and NK cell tumor infiltrates (Lo Presti, Hopps, & Caimi, 2017). Of note, the time span between exercise and tumor tissue sampling probably was too long to observe acute effects in this study. Keeping in mind that specifically CD56^dim^ NK cells are mobilized through exercise (figure 1D), further investigations may apply longer or more intense exercise regimes, which have been shown to provoke prolonged NK cell redistribution (Walsh et al., 2011).

Despite several analogies (NK cell mobilization, IL6 response) between the animal model and this first-in-human study, we did not detect any changes in NK cell tumor infiltrates. Several reasons may explain these discrepancies. Most importantly, researchers need to carefully differentiate between acute (single bouts) and chronic (regular bouts, also known as training) exercise.

Although acute effects on NK cell mobilization were highlighted in both the animal model and our study, our colleagues rather investigated chronic effects (six weeks of training) in regards to NK cell infiltrates in tumor tissue. On a systemic level chronic exercise does not alter NK cell numbers in humans(Valenzuela et al., 2022). Based on the findings of the current investigation, it is questionable whether acute exercise-induced NK cell mobilization and transient increased IL6 levels, permanently increase NK cell tumor infiltration.

In regards to systemic IL6 levels, acute and chronic exercise have opposite effects. As shown in the current investigation and in many other animal and human studies, acute exercise increases systemic IL6 levels in dependence of exercise type, duration and intensity, with most pronounced increases after intensive and prolonged aerobic exercise. In contrast, anti-inflammatory effects of chronic exercise mediate a decrease in resting levels of IL6 (Khalafi, Malandish, & Rosenkranz, 2021).

Apart from the discussion on acute and chronic effects of exercise, one should keep in mind that different tumor types were investigated. Although NK cell mobilization and redistribution is frequently sold as global mechanism mediating beneficial effects of exercise across various cancer types, this assumption has neither been tested nor been proven yet. Differences in immune accessibility are well described between various types of cancer. Therefore, our work investigating human tumor tissue for the first time in response to exercise should stimulate further research to evaluate whether physical exercise directly influences immune infiltration and possibly tumor growth and progress in different cancer types in humans. A more distinct focus on human tumor tissue is in progress (Holmen Olofsson et al., 2022) and urgently needed.

## Methods and Protocols

### Participant Recruitment and Informed Consent

The current study was approved by the ethics committee of the University Hospital Cologne and accords to the Declaration of Helsinki. The study was prospectively registered at the German Clinical Trials Register (DRKS00010442).

Twenty-two male patients with diagnosed high- or intermediate-risk prostate cancer (PSA > 10 ng/mL or Gleeson score ≥ 7 or cT2b) who were scheduled for radical prostatectomy (RP) were included in the study. Recruitment took place exclusively in the urology department of the University Hospital of Cologne. Patients were excluded if they had previously received radio- or chemotherapy, had experienced previous tumor diseases, as well as orthopedic, cardiovascular, or pulmonary disease precluded participation. After appropriate medical clarification, written informed consent was obtained from the participants surveyed.

### Interventions/ Exercise Training

The IG performed a single bout of endurance exercise on a bicycle ergometer (Ergoline) at moderate to vigorous intensity. The overall 30-minute endurance exercise included a 5-minute warm-up, followed by 20 minutes at a resistance on the ergometer equivalent to the resistance at 75% of the individual VO2peak during the baseline GXT test, and a 5-minute cool down. The CG stayed sedentary during the same period of time.

## METHODS

### Study Design and Randomization

The following study is based on a randomized controlled trial design. Initial contact was made by the urology department of the University Hospital of Cologne. A baseline examination with the performance of a graded exercise test (GTX) was conducted seven days prior to hospitalization. Using the maximal oxygen consumption as a stratification factor, the participants were randomized with concealed allocation into the intervention or control group using the minimization according to Pocock and Simon using the Randomization in Treatment Arms (RITA) software (Evidat, Sereetz, Germany).

Following the subjects’ completion of all medical examinations relevant to RP during the day, the intervention was performed on the evening before surgery at 7:00 p.m. in the urology department of the University Hospital of Cologne. Blood samples were taken before and two minutes after cessation of the bout of exercise.

### Anthropometrics and cardiorespiratory exercise test

A baseline test was performed seven days before the hospitalization of the participants. This included record of anthropometric data and determination of cardio-respiratory performance. For cardio-respiratory performance, a GXT was conducted on a stationary bicycle ergometer (Ergoline). During spiroergometry, measurements of maximal oxygen uptake (VO2_peak_) using METALYZER® 3B (CORTEX Biophysik GmbH, Leipzig, Germany), maximal heart rate (HRmax [beats per minute (bpm)]) using a heart rate sensor (Polar Electro GmbH Deutschland, Büttelborn, Germany) and maximal power output (PPO) was measured. Participants began with a five-minute warm-up at 20 W. The load was then increased by 20 W every two minutes. All participants were asked to maintain a cadence between 70 and 80 rpm. Exhaustion was defined when the respiratory quotient (VCO2/VO2) exceeded 1.1, decrease of cadence below 60 rpm, or subjective perceived exhaustion (Borg score > 18).

### Prostate cancer outcomes

Tumor pathology (TNM classification, ISUP grading) and clinical tumor characteristics (Prostate specific antigen, PSA) data were collected from medical records.

### Immunohistochemistry

Archived formalin-fixed, paraffin-embedded tumor tissue samples were sliced by the pathology department of the University Hospital Cologne and immunohistochemistry was performed. Subsequently, the slices were stained for NK cells (CD56) using mouse monoclonal antibody (123C3; dilution 1:500; Thermo Fisher Scientific, USA). All immunohistochemical stainings were performed using the Leica BOND-MAX stainer (Leica Biosystems, Germany) according to the protocol of the manufacturers. To determine NK cell infiltration, one researcher who was blinded to the experiment first scanned the entire sections at low magnification with a Zeiss Axiophot (KS300) microscope to assess the overall prevalence of infiltrating immune cells. Subsequently, two images of representative sections were taken. Images were recorded with a JENOPTIK GRYPHAX camera. One image was recorded with 20x magnification and without moving the sample. Another image was taken of the same area using 40x magnification. Finally, another blinded researcher evaluated stained areas (pixel/μm) using Image J software (1.51j8, Wayne Rasband, National Institutes of Health, USA).

### Blood Collection and Sample Preparation

Intravenous blood was drawn from the subjects at baseline (before exercise), immediately (2 minutes) and 12 hours after completing 30 minutes of exercise (IG) or after 30 minutes of sitting (CG). After clotting, serum was collected by centrifugation at 1100× g for 10 minutes. Afterwards, we aliquoted the serum and stored it at -80?C until sample measurement.

EDTA Blood samples were used for isolation of peripheral mononuclear blood lymphocytes (PBMCS). Blood samples were diluted with PBS (Thermo Fischer Scientific, USA) and carefully layered on top of a lymphocyte separation medium (Promo Cell, Germany). After centrifugation the PBMC containing layer were separated and washed with PBS. PBMCs were frozen in freezing medium (Thermo Fischer Scientific, USA) until analysis at -150°C.

### Assessment of inflammatory Markers

The inflammatory cytokines IL-6, IL10, TNF-α, TGF-β, MMP2, and MMP9 were measured following the manufacturer’s protocol using commercially available ELISA kits (R&D systems, Minneapolis, USA).

### Flow cytometry-based cytotoxicity assays

To measure tumor cell killing, thawed PBMCs were cultivated with prestained K562 tumor cells (tumor cells were provided by Prof. Carsten Watzel). Tumor cells were stained with cell tracker green (Thermo Fischer Scientific, USA) for 30 minutes. PBMCs were thawed and counted using an automated cell counter (OLS OMNI Life Science, Germany). PBMCs and tumor cells were mixed in E:T ratio of 1:1 and incubated for 4h at 37°C and 5% CO_2_. After incubation, cells were washed with PBS and stained with Zombie NIR™ viability stain (Biolegend, USA). Thereafter, cells were stained with anti-CD3 PE, anti-CD56 BV421 and anti-CD16 AF647 (Biolegend, USA) and fixated using a paraformaldehyde containing Fixation buffer (Biolegend, USA).

Cell analysis was performed using a BD LSRFortessa™ Cell Analyser (BD Bioscience, Germany) and cells were gated as cell tracker green positive tumor cells and were divided in vital and dead cells via Zombie NIR™. Furthermore, NK-cells were characterized as CD3^+^CD56^+^ and divided in CD56^bright^ (CD56^++^CD16) and CD56^dim^ (CD56^+^ CD16^-/+^) (Montaldo et al., 2013). Changes in lysis activity were adjusted for exercise driven changes in CD56^dim^/CD56^bright^ ratio.

### Statistical Analysis

All statistical analyses were performed by a blinded investigator. Outcome data were first tested for normality. In case of violation, a log transformation was conducted to remove the skewness of raw data. We tested our sample for possible baseline differences between groups in anthropometric and performance characteristics using independent t-, and chi square tests. Intervention effects on all outcomes were assessed using baseline (t1) adjusted analysis of co-variance (ANCOVA) for repeated measures (factor time: t1, t2, t3; factor group: IG vs CG). In case of significant time and interaction effects, Bonferroni corrected simple effects analysis were performed. Acute changes (t1, t2) in NK cell activity and Immune cell infiltrates in tumor tissues were compared using one-way ANCOVA. Exercise induced alterations of lytic activity were adjusted for changes (t1, t2) in CD56^dim^/CD56^bright^ ratio, while circulating NK cells at t3 were put as co variate for tumor infiltrates. To evaluate the associations between circulating NK cells, (anti)inflammatory markers immediately prior to surgery and immune cell infiltration correlation analyses were performed. The level of significance is set at 5%. The level of significance was set at p ≤ 0.05. All statistical analyses were conducted using IBM SPSS Statistics (version 28).

## Data Availability

All data produced in the present study are available upon reasonable request to the authors.

## Resource availability

### Lead contact

Further information and requests for resources and reagents should be directed to and will be fulfilled by the lead contact, Philipp Zimmer

### Material Availability

This study did not generate new unique reagents.

### Data and Code Availability

All data reported in this paper will be shared by the lead contact upon request. This paper does not report original code. Any additional information required to reanalyze the data reported in this paper is available from the lead contact upon request.

## REFERENCES

Chow, L. S., Gerszten, R. E., Taylor, J. M., Pedersen, B. K., van Praag, H., Trappe, S., … Snyder, M. P. (2022). Exerkines in health, resilience and disease. Nat Rev Endocrinol. doi:10.1038/s41574-022-00641-2

Eschke, R. K., Lampit, A., Schenk, A., Javelle, F., Steindorf, K., Diel, P., … Zimmer, P. (2019). Impact of Physical Exercise on Growth and Progression of Cancer in Rodents-A Systematic Review and Meta-Analysis. Front Oncol, 9, 35. doi:10.3389/fonc.2019.00035

Holmen Olofsson, G., Mikkelsen, M. K., Ragle, A. M., Christiansen, A. B., Olsen, A. P., Heide-Ottosen, L., Thor Straten, P. (2022). High Intensity Aerobic exercise training and Immune cell Mobilization in patients with lung cancer (HI AIM)-a randomized controlled trial. BMC Cancer, 22(1), 246. doi:10.1186/s12885-022-09349-y

Joisten, N., Schenk, A., & Zimmer, P. (2020). Talking About Physical “Activity” or “Inactivity”? The Need of Accurate Activity Controlling in Exercise Studies in Rodents. Front Physiol, 11, 611193. doi:10.3389/fphys.2020.611193

Khalafi, M., Malandish, A., & Rosenkranz, S. K. (2021). The impact of exercise training on inflammatory markers in postmenopausal women: A systemic review and meta-analysis. Exp Gerontol, 150, 111398. doi:10.1016/j.exger.2021.111398

Lo Presti, R., Hopps, E., & Caimi, G. (2017). Gelatinases and physical exercise: A systematic review of evidence from human studies. Medicine (Baltimore), 96(37), e8072. doi:10.1097/md.0000000000008072

Montaldo, E., Del Zotto, G., Della Chiesa, M., Mingari, M. C., Moretta, A., De Maria, A., & Moretta, L. (2013). Human NK cell receptors/markers: a tool to analyze NK cell development, subsets and function. Cytometry A, 83(8), 702–713. doi:10.1002/cyto.a.22302

Orange, S. T., Jordan, A. R., & Saxton, J. M. (2020). The serological responses to acute exercise in humans reduce cancer cell growth in vitro: A systematic review and meta-analysis. Physiol Rep, 8(22), e14635. doi:10.14814/phy2.14635

Patel, A. V., Friedenreich, C. M., Moore, S. C., Hayes, S. C., Silver, J. K., Campbell, K. L., … Matthews, C. E. (2019). American College of Sports Medicine Roundtable Report on Physical Activity, Sedentary Behavior, and Cancer Prevention and Control. Med Sci Sports Exerc, 51(11), 2391–2402. doi:10.1249/mss.0000000000002117

Pedersen, L., Idorn, M., Olofsson, G. H., Lauenborg, B., Nookaew, I., Hansen, R. H., … Hojman, P. (2016). Voluntary Running Suppresses Tumor Growth through Epinephrine-and IL-6-Dependent NK Cell Mobilization and Redistribution. Cell Metab, 23(3), 554–562. doi:10.1016/j.cmet.2016.01.011

Rumpf, C., Proschinger, S., Schenk, A., Bloch, W., Lampit, A., Javelle, F., & Zimmer, P. (2021). The Effect of Acute Physical Exercise on NK-Cell Cytolytic Activity: A Systematic Review and Meta-Analysis. Sports Med, 51(3), 519–530. doi:10.1007/s40279-020-01402-9

Schlagheck, M. L., Walzik, D., Joisten, N., Koliamitra, C., Hardt, L., Metcalfe, A. J., … Zimmer, P. (2020). Cellular immune response to acute exercise: Comparison of endurance and resistance exercise. Eur J Haematol, 105(1), 75–84. doi:10.1111/ejh.13412

Schmitz, K. H., Campbell, A. M., Stuiver, M. M., Pinto, B. M., Schwartz, A. L., Morris, G. S., … Matthews, C. E. (2019). Exercise is medicine in oncology: Engaging clinicians to help patients move through cancer. CA Cancer J Clin, 69(6), 468–484. doi:10.3322/caac.21579

Valenzuela, P. L., Saco-Ledo, G., Santos-Lozano, A., Morales, J. S., Castillo-García, A., Simpson, R. J., … Fiuza-Luces, C. (2022). Exercise Training and Natural Killer Cells in Cancer Survivors: Current Evidence and Research Gaps Based on a Systematic Review and Meta-analysis. Sports Med Open, 8(1), 36. doi:10.1186/s40798-022-00419-w

Walsh, N. P., Gleeson, M., Shephard, R. J., Gleeson, M., Woods, J. A., Bishop, N. C., … Simon, P. (2011). Position statement. Part one: Immune function and exercise. Exerc Immunol Rev, 17, 6–63.

